# Progression of COVID-19 in Indian States - Forecasting Endpoints Using SIR and Logistic Growth Models

**DOI:** 10.1101/2020.05.15.20103028

**Authors:** Bhoomika Malhotra, Vishesh Kashyap

**Affiliations:** Department of Mathematics Kamala Nehru College University of Delhi Delhi, India; Department of Mechanical, Production & Industrial and Automobile Engineering Delhi Technological University Delhi, India

## Abstract

COVID-19 has led to the most widespread public health crisis in recent history. The first case of the disease was detected in India on 31 January 2019, and confirmed cases stand at 74,281 as of 13 May 2020. Mathematical modeling can be utilized to forecast the final numbers as well as the endpoint of the disease in India and its states, as well as assess the impact of social distancing measures. In the present work, the Susceptible-Infected-Recovered (SIR) model and the Logistic Growth model have been implemented to predict the endpoint of COVID-19 in India as well as three states accounting for over 55% of the total cases – Maharashtra, Gujarat and Delhi. The results using the SIR model indicate that the disease will reach an endpoint in India on 12 September, while Maharashtra, Gujarat and Delhi will reach endpoints on 20 August, 30 July and 9 September respectively. Using the Logistic Regression model, the endpoint for India is predicted on 23 July, while that for Maharashtra, Gujarat and Delhi is 5 July, 23 June and 10 August respectively. It is also observed that the case numbers predicted by the SIR model are greater than those for the Logistic Growth model in each case. The results suggest that the lockdown enacted by the Government of India has had only a moderate impact on the spread of COVID-19, and emphasize the need for firm implementation of social distancing guidelines.

## 1 Introduction

The severe acute respiratory syndrome coronavirus 2 (SARS-nCoV-2) or 2019 novel coronavirus (2019-nCoV) has emerged as a significant worldwide health threat over the initial months of 2020.The Coronavirus Disease (COVID-19) is a highly contagious infectious respiratory disease caused by this coronavirus, which belongs to a family of viruses responsible for communicable respiratory disorders such as Severe Acute Respiratory Syndrome (SARS) and Middle East Respiratory Syndrome (MERS) [1]. The most common modes of transmission of COVID-19 are through contact and respiratory droplets [2]. As of 13 May 2020, there have been 4,139,794 cases of COVID-19 globally, resulting in 285,328 deaths [3].

The 2019-nCoV virus originated between October and December 2019, with bats as the natural reservoir host [4]. However, transmission from bats to humans was likely through an intermediary host [5]. On 31 December 2019, the Wuhan Municipal Health Commission reported a cluster of 27 pneumonia cases of unknown origin in Wuhan, Hubei province, China. A number of the initial cases were found to have common contact with the Huanan Seafood Wholesale Market[6]. On 7 January 2020, the virus was identified as a novel coronavirus, and its genetic sequence was shared on 11 January 2020. On the same day, the first fatality from COVID-19 was reported in Wuhan [7].

The first case of COVID-19 outside China was reported on 13 January 2020 in Thailand [8]. U.S.A. reported the first travel-related COVID-19 case on 15 January 2020 [9]. A day later, Japan’s first COVID –19 case was reported [10]. By the end of January 2020, cases had been reported in South Korea, Taiwan, Hong Kong, Macau, Singapore, Vietnam, France, Nepal, Australia, Canada, Malaysia, Cambodia, Germany, Sri Lanka, Finland, United Arab Emirates, India, Italy, Philippines, Russia, Spain, Sweden and the United Kingdom, with a total of 9,826 confirmed cases – 9,720 inside China and 106 in other countries [11]. On 30 January 2020, the World Health Organization declared the 2019-nCoV outbreak a Public Health Emergency of International Concern [12].

The first COVID-19 positive case in India was reported on 30 January 2020 in a returnee from Wuhan. Two similar cases were reported on 2 and 3 February, and all 3 were reported recovered on 14 February 2020 [13]. On 2 March 2020, 2 travel-related COVID-19 cases were reported in Delhi and Hyderabad [14]. On 4 March, 15 Italian tourists with a travel history across Rajasthan tested positive for COVID-19 in Delhi [15]. In response to the outbreak screening of all international passengers was initiated on 6 March 2020, and all non-essential traveller visas were suspended on 13 March 2020 [16]. A three-week nationwide lockdown was imposed from 25 March to 14 April [17], which was later extended to 3 May and then to 17 May. The total number of cases in India reached 1,000 on 28 March and 10,000 on 14 April. The total number of confirmed cases in India as of 13 May 2020 stands at 74,281, with 24,385 recoveries and 2,415 deaths [18]. The case fatality ratio (CFR) for India increased from 1.9% on 15 March 2020 to 3.6% on 12 April 2020 [19], and stands at 3.2% (as of 13 May 2020).

Mathematical modeling is regularly used in order to analyze the dynamics of the propagation of diseases and predict future transmission trends. A number of studies using mathematical modeling have been performed for COVID-19, a significant majority focusing on transmission in China, Japan and the Diamond Princess cruise ship, where the virus spread initially. The reproduction rate of a disease (*R*_0_) represents the number of persons infected by each infected person. Liu et al. [20] reviewed studies which attempted to calculate the *R*_0_ of COVID-19 in China, and observed that studies which used mathematical modeling reported an average *R*_0_ of 4.2, while those that used statistical methods reported an average *R*_0_ of 2.67. Both these values were observed to be higher than the WHO estimate of 1.95. Zhao et al. [21] used the Poisson process approach to estimate the *R*_0_ for the Diamond Princess cruise ship at 2.1. Tindale et al. [22] statistically estimated *R*_0_ for Singapore at 1.97 and Tianjin, China at 1.87. They also concluded that the mean incubation period for COVID-19 was significantly greater than the serial interval, raising the possibility of asymptomatic transmission. Similar conclusions were also obtained by Mizumoto et al. [23] for the Diamond Princess cruise ship with an asymptomatic ratio of 17.9%, Nishiura et al.[24, 25] for Japan with an asymptomatic ratio of 41.6% and Zhao et al. [26] for Hong Kong.

The Logistic Growth model and the susceptible-infected-recovered (SIR) model are among the most frequently used for epidemiological prediction, and have also been applied in the case of the COVID-19 pandemic. Roosa et al. [27] used a generalized logistic growth model to assess the impact of containment strategies in China and predict final case numbers. Vattay [28] implemented the logistic growth model to analyze the similarity between trends of death numbers in Hubei, China and Italy, and predicted an end date for further growth of COVID-19 in Italy. Wu et al. [29] analyzed the growth of COVID-19 in individual Chinese provinces using 4 logistic growth models, and further implemented them to predict case sizes in other countries of Europe, the Americas and Asia. Zhou et al. [30] forecasted the spread of COVID-19 using the logistic growth model and the basic SIER model, and observed that the pandemic size reported using the logistic model was considerably smaller. Batista [31] estimated the eventual size of COVID-19 cases worldwide using the logistic regression model as well as the SIR model, and observed negligible (<1%) difference among the forecasts of the two. Using observations from China and South Korea as benchmarks, Gaeta [32] analyzed the progression of COVID-19 in Italy using the SIR model and observed the initial growth rate in the countries to be similar in nature, drawing conclusions on the impact of future restrictive measures. Calafiore et al. [33] developed a modified SIR model for the forecast of COVID-19 numbers in individual Italian provinces, which contained an additional proportionality factor comparing the confirmed cases and the actual infected number. Ku et al. [34] utilized the modified Bass-SIR model to forecast the progression of the disease in Wuhan, predicting a second wave of cases if restrictions were lifted. Wangping et al. [35] employed an extended SIR model to estimate the basic reproduction number for Hunan, China and Italy, and forecast progression of COVID-19. They estimated *R*_0_ to be 4.34 for Italy an 3.15 for Hunan, forecasting an endpoint for the disease.

The implementation of mathematical modeling for the forecast of COVID-19 progression in India has been limited as compared to the aforementioned countries. Pandey et al. [1] utilized the SEIR and regression models to predict the progress of COVID-19 in India over two weeks, and observed that the regression model yielded a number greater by 15.7%. Dhanwant and Ramanathan [36] utilized the SIR model to predict the growth of COVID-19 during the initial 3-week lockdown, and if the lockdown were lifted on 14 April, concluding that the extent of social distancing was not sufficient to stem the growth of the disease. Poonia and Azad [37] analyzed the progress of COVID-19 in individual Indian states, forecasting regions where restrictions may be relaxed post-April. Sardar et al. [38] utilized the SEIR model to forecast the impact of the initial lockdown on the spread of COVID-19, concluding that the same would be limited unless the restrictions were extended.

Given the considerable restrictions applied on inter-state travel in India, it way be deduced that the progress of COVID-19 in India will be highly state-specific. However, through a review of previous literature, it is observed that work on forecasting the spread of the disease in specific states is limited. The present work aims to utilize the SIR and Logistic Growth models to predict future spread of COVID-19 in India and the three highest-impacted states of India-Maharashtra, Gujarat and Delhi.

## 2 Methodology

Information regarding the number of confirmed infections and deaths was retrieved from the website *https://www.mygov.in/covid-19*. The current work aims to draw a comparison and analyse the spread of SARSCoV-2 virus in the aforementioned states by employing two models; The Susceptible-Infected-Recovered (SIR) Model and the Logistic Growth Model. Modeling was initiated after 100 confirmed cases had been reported in each individual state in order to avoid significant initial variations in data.

### 2.1 The Susceptible-Infected-Recovered (SIR) Model

The SIR model compartmentalises the entire population(N) into 3 categories; Susceptible (S), Infectious (I) and Recovered (R). Parameters *β* and *γ* are used to formulate the following differential equations to model the flow of individuals through these compartments:

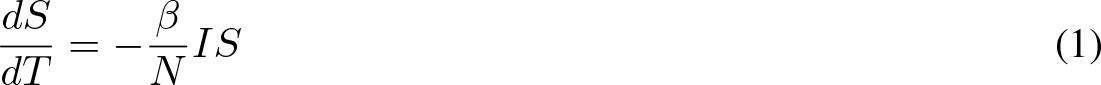

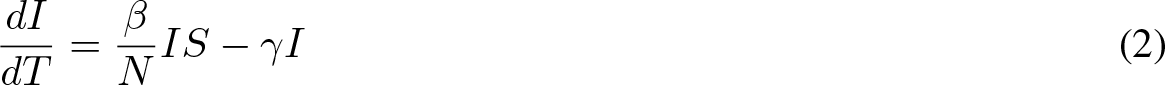

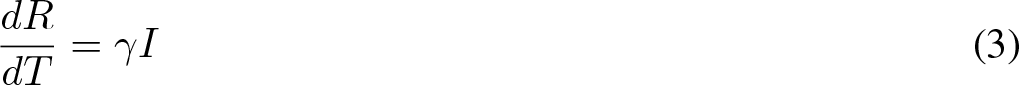

#### Model States and Parameters

- *t* is the daily time-parameter
- *S*(*t*) denotes the total susceptible population at time *t*
- *I*(*t*) denotes the number of active infections at time *t*
- *R*(*t*) denotes the total number of recoveries and deaths at time *t*
- *β* denotes the contact rate which factors the rise of infections due to interactions between the susceptible and infected population. This parameter takes into consideration the population size, reproduction number *R*_0_ and exposure-factor.
- 1*/γ* denotes the average infectious period i.e the rate at which infections are eliminated either due to recovery or demise. We assume that the recovered individuals would not spread the infection again.
- *N* denotes the total population where *S* + *I* + *R* = *N*
- *R*_0_ is the basic reproduction number which is defined as the expected number of secondary cases produced by a single infection in a susceptible population. It may be noted that *β* = *R*_0_*γ*.

Eliminating *I* from equations (1) and (3), and applying limit *t → ∞*, we obtain

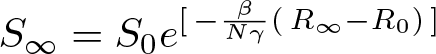

where *S*_0_ and *R*_0_ represent the initial conditions *S*(0) and *R*(0), respectively, and *S_∞_* represents the final susceptible population.

As the final number of infections tends to zero, the final number of recovered individuals, *R_∞_*, becomes

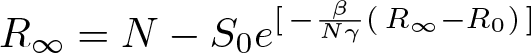

since

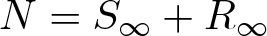

We now proceed by estimating the parameters *β* and *γ*, and the initial values by minimizing the difference between confirmed and predicted number of cases.

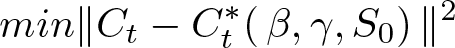

where *C_t_* = (*C*_1_*, C*_2_*, … C_n_*) represents the confirmed number of cases and 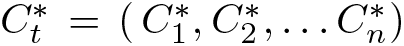 denotes the corresponding predictions calculated by the model at time *t_i_, i ∈* 1 *… n*.

### 2.2 The Logistic Growth Model

The Logistic Growth Model suggests that the rate of growth decreases as the output approaches the model’s upper bound, called the carrying capacity. In terms of COVID-19, the relative growth rate decreases when the number of infections is approaching the final size of the epidemic. In other words, the rate of change in the number of new cases per capita linearly decreases with the number of cases. The model is expressed by the following differential equation:

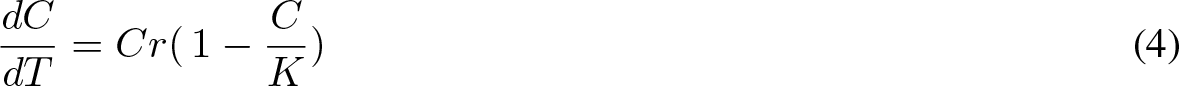

#### Model States and Parameters

- *t* is the daily time-parameter
- *C*(*t*) denotes the total number of confirmed infections at time *t*
- *r* denotes the infection rate which sets the typical time scale of the growth process of the epidemic. This parameter takes into account interaction, restrictive measures, tests being conducted for detection and exposure (elements contributing towards change in the number of cases). *r* is also responsible for the steepness of the graph.
- *K* denotes the final epidemic size, also known as the ‘carrying capacity’.

From equation (4), we obtain,

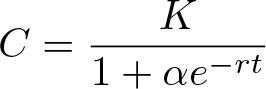

where *C*_0_ represents the initial condition *C*(0), and *α* represents a constant equivalent to 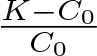.

The growth rate peaks at time *t_p_* when the condition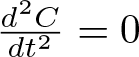, i.e.

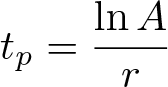

During this peak time, number of cases is equivalent to 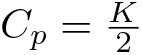 and growth rate is 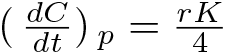.

Further, the doubling time is

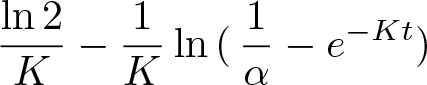

The following regression model can be used to estimate the parameters of the Logistic Growth Model,

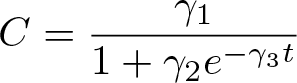

when *γ*_1_*, γ*_2_*, γ*_3_ are dependent upon the available data.

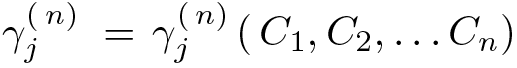, *j* ∈ {1, 2, 3} perfectly captures the time dependency of the parameters where *C_i_* denotes the total infections at time *t_i_, i ∈* 1 *… n*.

In accordance with our assumption, *γ_i_*(*t*) will converge to a finite value as the epidemic reaches its carrying capacity.

## 3 Results and Discussion

The SIR and Logistic Growth models have been implemented based on the trends in COVID-19 cases obtained for all of India as well as the highly affected states of Maharashtra, Gujarat and Delhi until 13 May. MATLAB functions have been utilized in order to forecast future trends. A high goodness of fit (*R*^2^> 0.99, p < 0.0001) was obtained for all the cases and models studied.

The total number of confirmed COVID-19 cases in India reached 74,281 on 13 May with the highest surge recorded at 4,213 cases on 11 May. The SIR model (Figure 1) forecasts the attainment of peak on 26 May with 4,913 (95% CI, 4,700–5,126) cases. The model also predicts a total of 277,858 (95% CI, 274,081–261,635) cases by 12 September. With an estimated *R*_0_ of 1.21, the SIR model’s trajectory suggests a decline in the number of cases almost 10 days after the cessation of Lockdown 3. The Logistic Growth model (Figure 2) predicts the crest will be attained on 16 May at 3,503 (95% CI, 3,247–3,759) cases. It further converges to a grand total of 153,513 (95% CI, 150,101–156,925) cases with an expected endpoint of 23 July. The model also depicts that deceleration will initiate towards the end of the third phase of the lockdown indicating a slim reduction in the number of cases because of the restrictive measures imposed.

**Figure 1:**
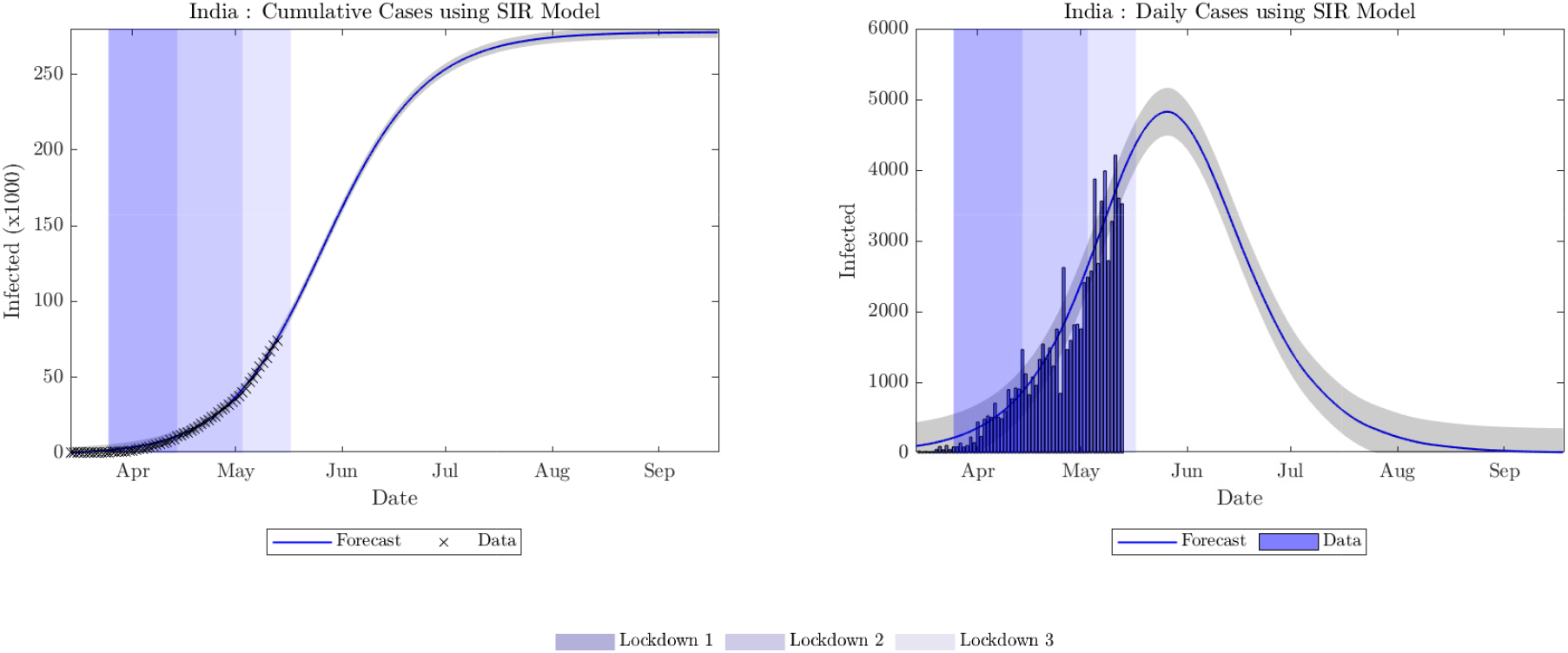
Cumulative and daily cases forecast for India using the SIR model.

**Figure 2:**
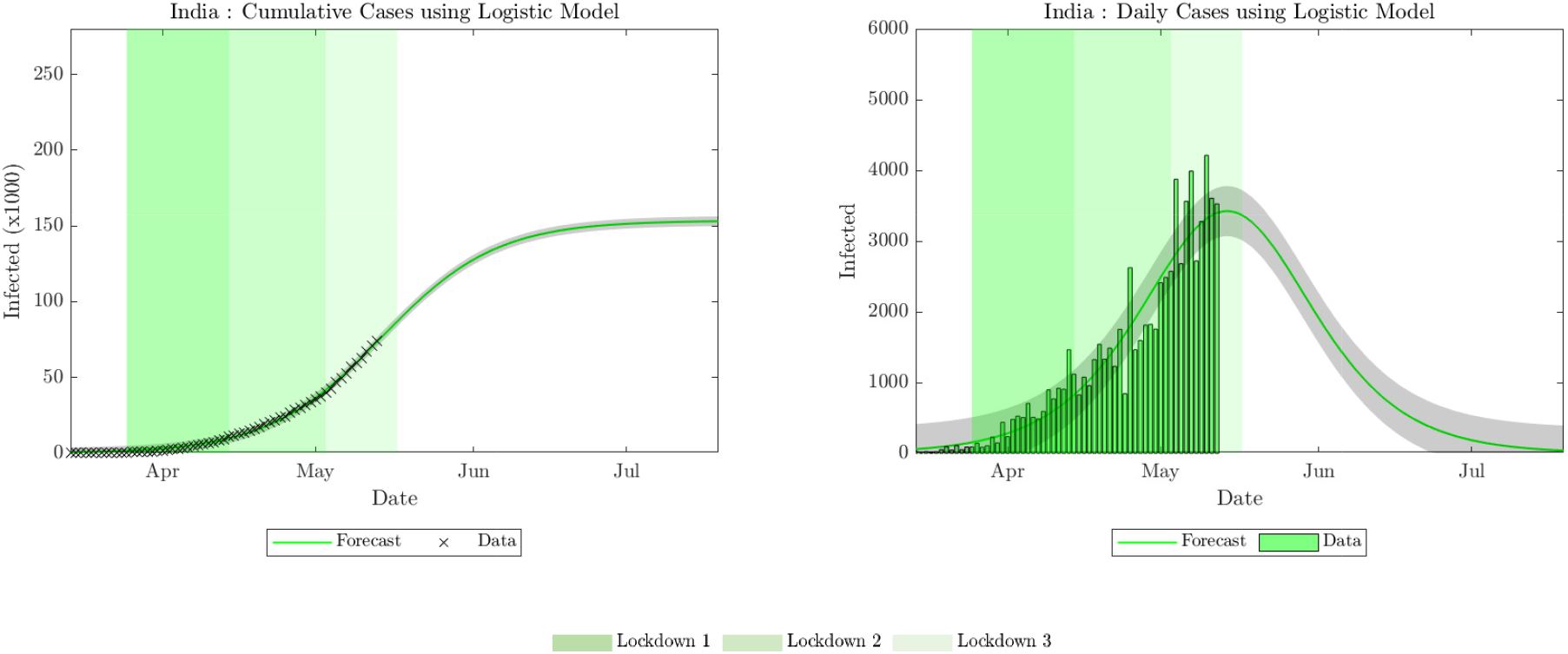
Cumulative and daily cases forecast for India using the Logistic Growth model.

Maharashtra recorded 24,427 confirmed cases of COVID-19 till 13 May 2020 with a maximum increase of 1,943 cases being observed on 11 May. The estimated *R*_0_ for the state is 1.20. The trajectory of the SIR model (Figure 3) indicates that the peak of the pandemic is likely to be attained on 20 May with an estimated 1,578 cases (95% CI, 1,426–1,730). The model also predicts an endpoint of 20 August with a total of 70,917 (95% CI, 70,275–71,559) cases. The Logistic Growth model (Figure 4) converges at 50,874 (95% CI, 50,453–51,295) cases on 5 July with the peak being attained on 15 May with 1,327 (95% CI, 1156–1498) cases. It is interesting to observe that the logistic model suggests a decline in the number of cases towards the end of the third stage of the lockdown, rendering it effective, whilst the SIR model depicts that cases will peak well after the lockdown is over, implying the need for further restrictive measures.

**Figure 3:**
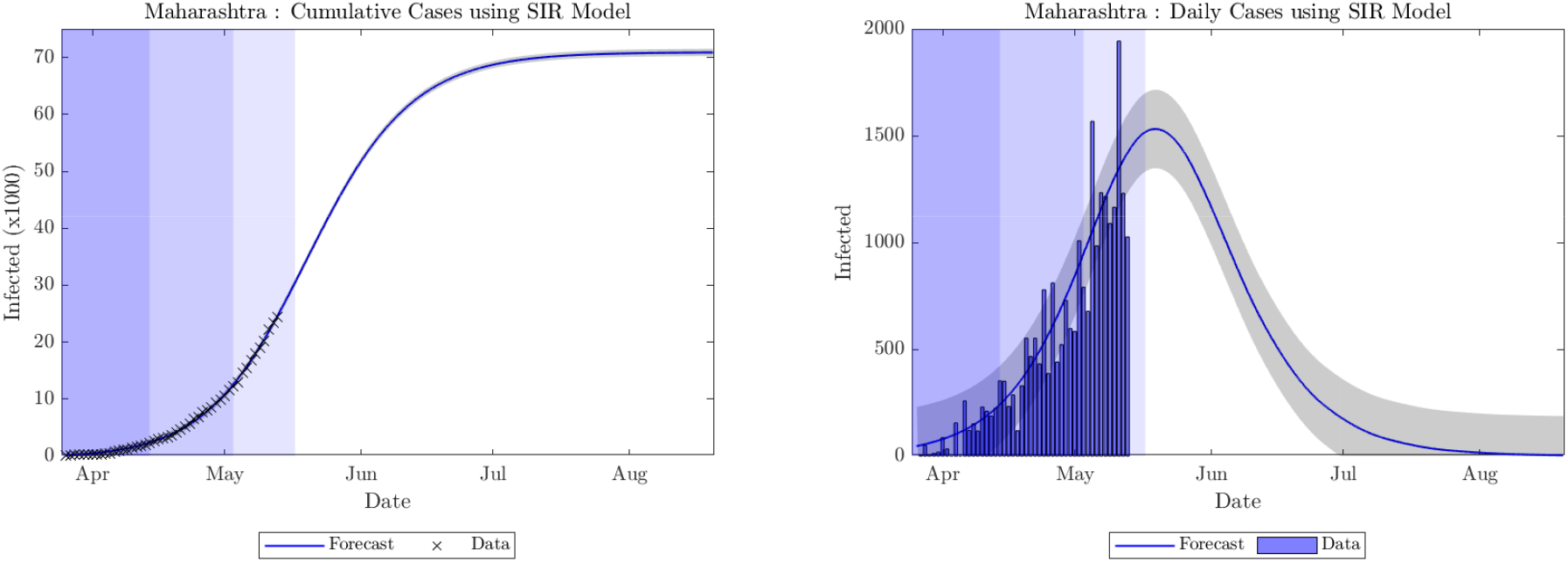
Cumulative and daily cases forecast for Maharashtra using the SIR model.

**Figure 4:**
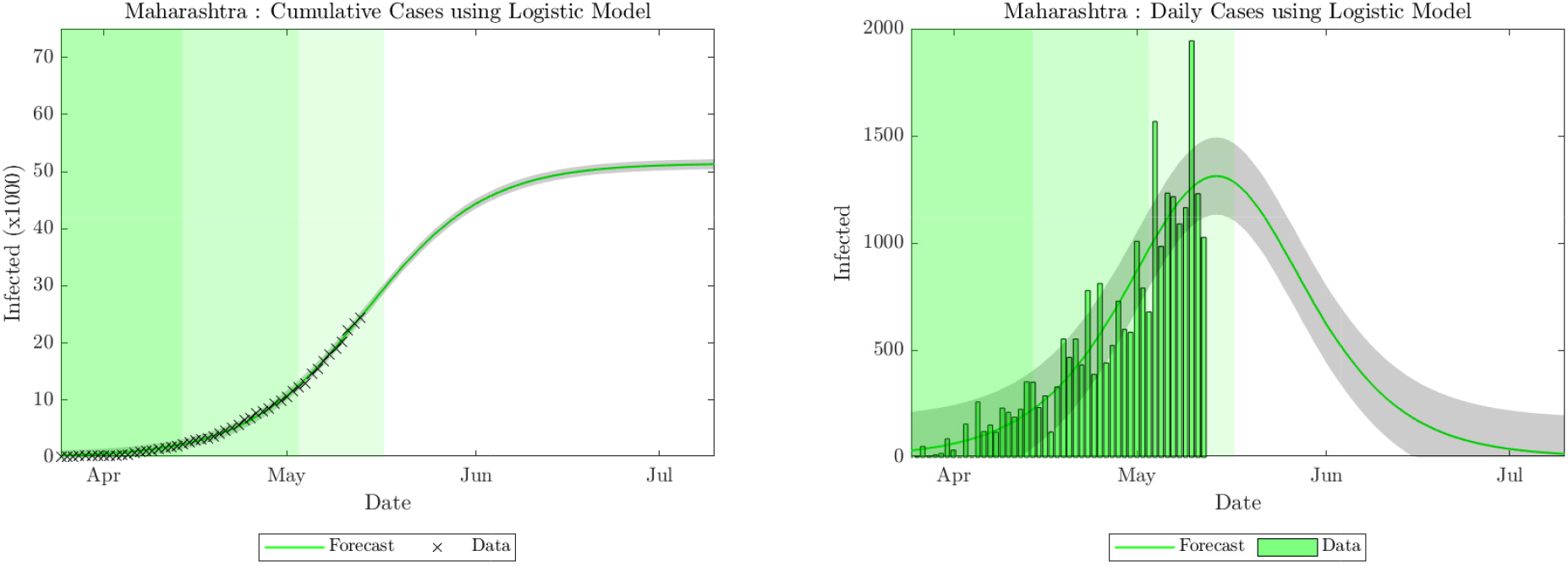
Cumulative and daily cases forecast for Maharashtra using the Logistic Growth model.

Gujarat has recorded 8,903 cases as of 13 May, the second highest tally in India after Maharashtra. The highest hike in cases recorded in a span of 24 hours was reported to be 441 cases on 6 May. The SIR model (Figure 5) suggests that cases in Gujarat will begin to decline after attaining a peak at 373 (95% CI, 313–433) cases on 11 May, which is in agreement with the actual data. The total number of cases is expected to reach 16,011 (95% CI, 15,561–16,461) by 30 July. On the other hand, the logistic model (Figure 6) converges to 12,114 (95% CI, 11,540–12,688) cases with an endpoint of 23 June. The model also implies that epidemic has already peaked at 362 (95% CI, 314–410) cases on 6 May. The estimated *R*_0_ value for Gujarat is 1.20. The predictions of both models are in agreement regarding the declining of cases in the third phase of the lockdown, which is in accordance with the actual data.

**Figure 5:**
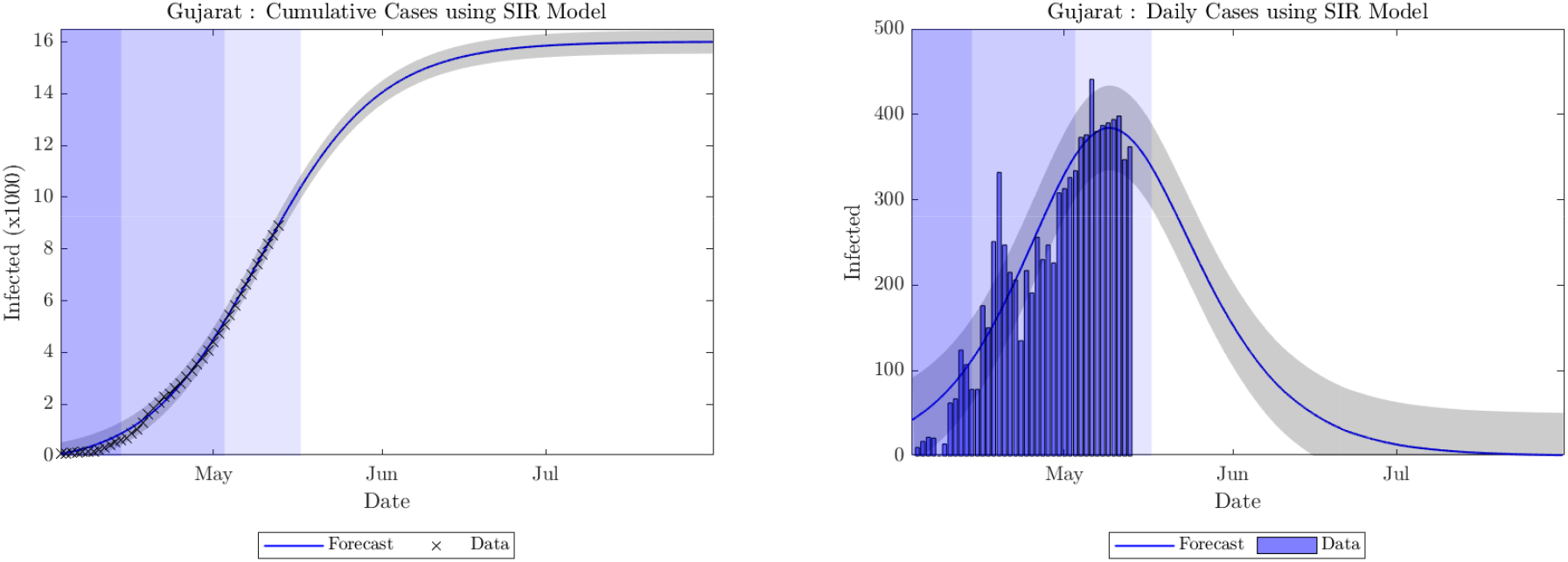
Cumulative and daily cases forecast for Gujarat using the SIR model.

**Figure 6:**
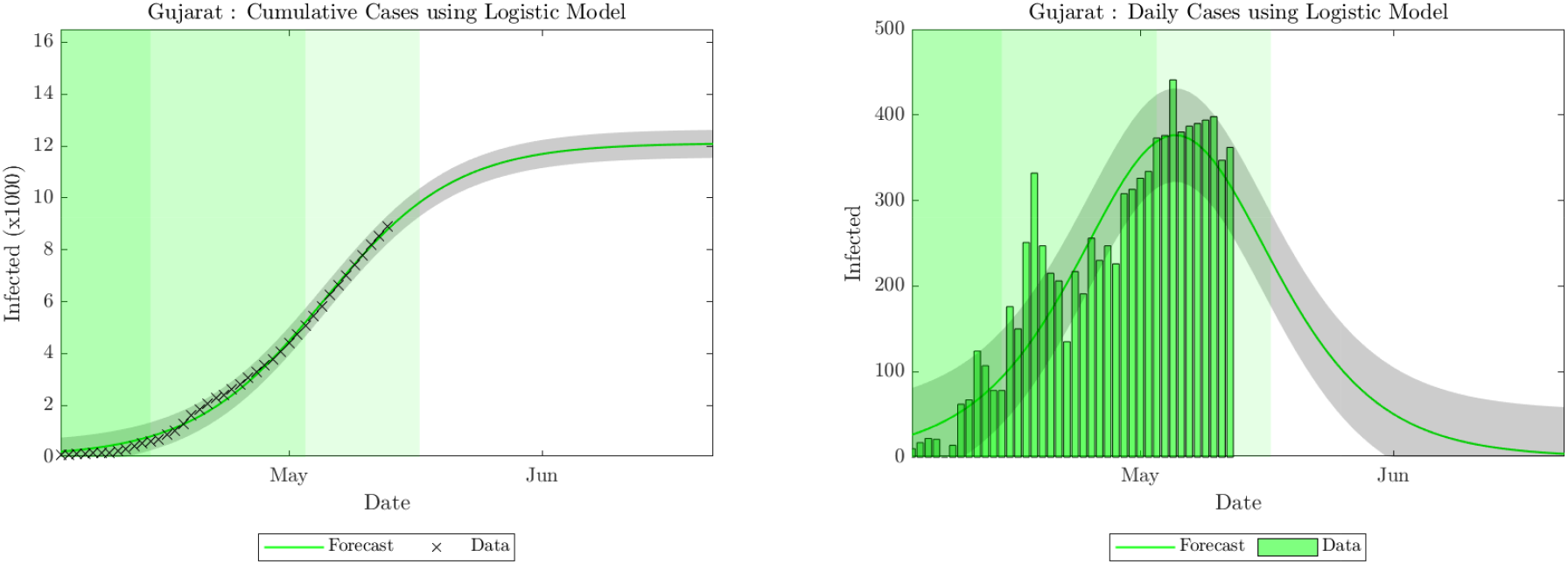
Cumulative and daily cases forecast for Gujarat using the Logistic Growth model.

Delhi amounts for 7,639 or 10.28% of the total 74,281 confirmed cases of COVID-19 in India. A maximum increase of 406 cases was recorded on 13 May. The SIR model (Figure 7) predicts a total of 40,002 (95% CI, 39,252–40,752) cases by 9 September. A crest is predicted on 4 July with 653 (95% CI, 547–759) cases. The Logistic Growth model (Figure 8) forecasts 29,387 (95% CI, 29,239–30,129) cases with an endpoint of 10 August. The attainment of the peak is expected on 25 June at 521 (95% CI, 425–617) cases. With an estimated *R*_0_ value of 1.13, the trajectories of both the models depict a deceleration of the curve after the third phase of the lockdown ends.

**Figure 7:**
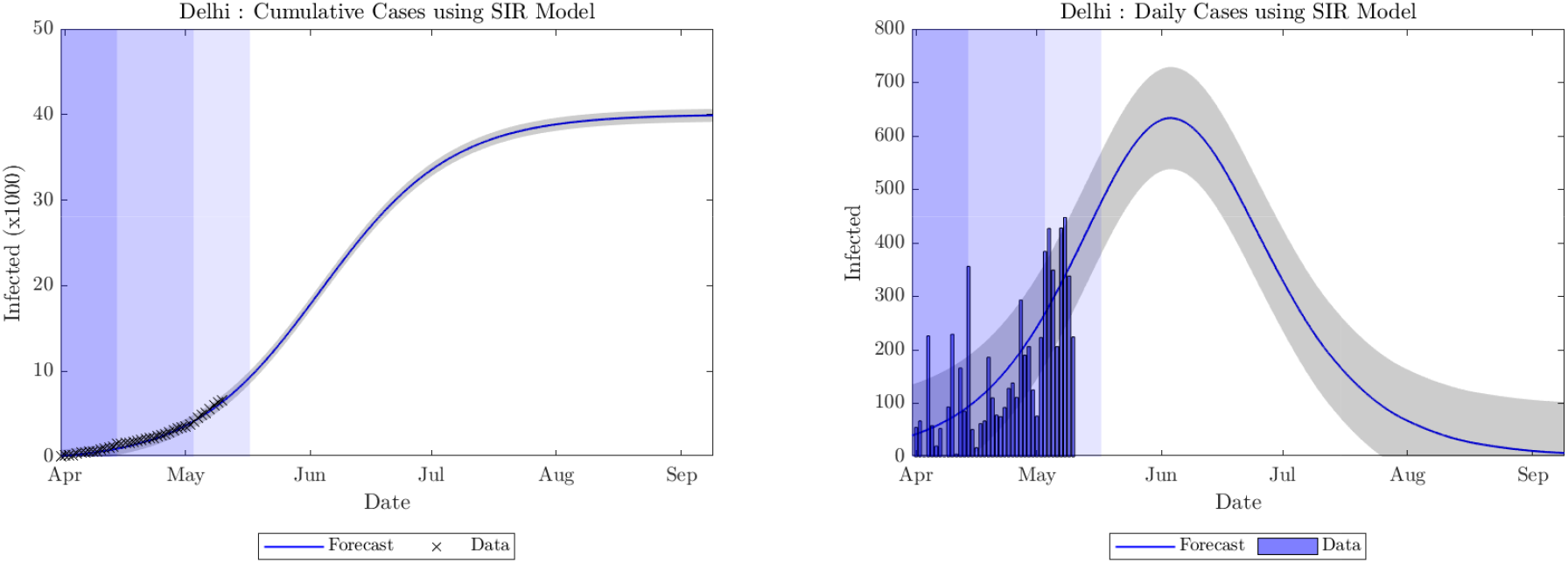
Cumulative and daily cases forecast for Delhi using the SIR model.

**Figure 8:**
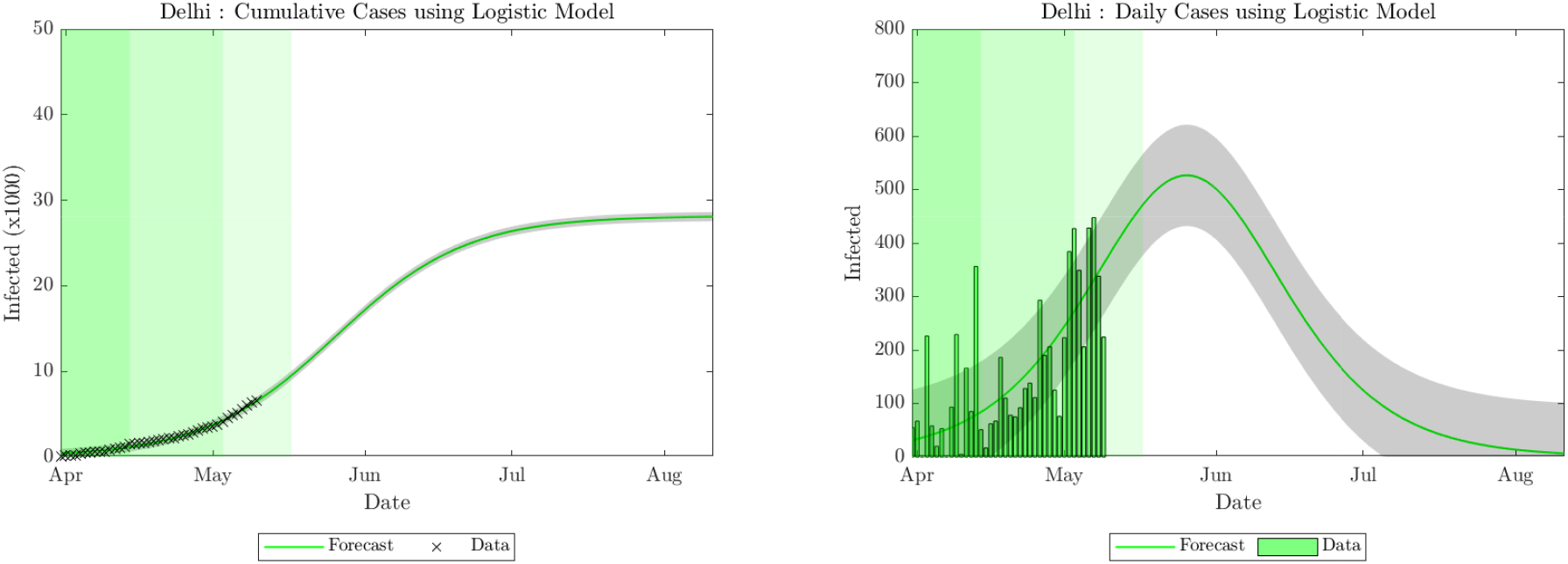
Cumulative and daily cases forecast for Delhi using the Logistic Growth model.

On an analysis of the results obtained, a number of deductions may be made from the trends yielded by both models. The most prominent observation deals with the region-specific nature of the propagation of the pandemic. The endpoints of the pandemic vary widely with region, with a cessation in cases being forecast for 30 July in Gujarat and 9 September in Delhi by the SIR model. The logistic model similarly forecasts wide variation in endpoints, from 23 June in Gujarat to 10 August in Delhi. A possible reason for this may be the restriction on inter-state travel implemented in India since 25 March 2020, which seem to have stemmed the propagation of the disease among states. Similar results were obtained by Calafiore et al. [33], who predicted varying trends for the spread of COVID-19 in different Italian provinces.

At the same time, the SIR model indicates evident similarities in the propagation of COVID-19 in all the regions studied. The contact rate *β* is observed to be similar (0.54–0.56) for all the three states studied (Table 1). This indicates a partial success of social distancing measures, suggesting that the rate of transmission of disease is similar in all highly-affected regions irrespective of demographic indices such as population density. The value of average infectious period 1/*γ* is also observed to be minimally variant among the states studied, with a minimum of 2.04 for Delhi and a maximum of 2.22 for Maharashtra. The value of *β* for India is 0.42, indicating the diminishing impact of other, less-affected regions, while 1/*γ* is 2.94, which is similar to the median incubation period of 3 days observed in China [39]. The estimated value of *R*_0_ for India as a whole is minimally variant from the *R*_0_ values of individual Indian states which constitute 55% of the total cases nationwide (Table 1), with a maximum variation of 6.6% for Delhi. These values are also in concordance with reported values [40]. The Logistic Growth model forecasts lower case numbers as well as early endpoints for all the regions studied. It is also observed to be significantly less sensitive to variations in recent case numbers as compared to the SIR model. Using the logistic model, significant variations in the infection rate parameter *r* may be observed among states. The lowest infection rate of 0.07 is estimated for Delhi while the highest is estimated for Gujarat (0.12). The forecast of the logistic model indicates that the impact of restrictive measures has varied widely over the country, and demographic factors have a role to play in determining the progression of disease in different states.

**Table 1:** Estimated values.

|  | SIR |  |  | Logistic Growth |  |  |
| --- | --- | --- | --- | --- | --- | --- |
| | $\beta$ | $\gamma$ | $R_0$ | $K$ | $\alpha$ | $r$ |
| India | 0.42 | 0.34 | 1.21 | 153513.79 | 231.36 | 0.09 |
| Maharashtra | 0.54 | 0.45 | 1.20 | 51438.49 | 165.08 | 0.10 |
| Gujarat | 0.56 | 0.47 | 1.20 | 12114.91 | 55.29 | 0.12 |
| Delhi | 0.55 | 0.49 | 1.13 | 28170.64 | 65.62 | 0.07 |

Significant deductions may also be drawn from the forecasts obtained regarding the impact of restrictive measures such as the lockdown on the propagation of COVID-19 in India. In the states of Gujarat and Maharashtra as well as for India as a whole, the progressively-eased lockdown totaling 53 days seems to have had a prominent impact, with the SIR model forecasting a peak in cases being reached inside the lockdown period for Gujarat and within 15 days of Lockdown 3 for India and Maharashtra. The logistic model forecasts that a peak in cases will be reached by Maharashtra and Gujarat as well as India within the lockdown period. Both the models indicate that the impact of the lockdown in Delhi has been nominal, with a peak being reached 15 to 30 days after Lockdown 3 as per existing trends. Previous studies such as those by Sardar et al. [38] and Singh and Adhikari [41] forecast that a complete lockdown would lead to a sharp drop in cases during the initial 21-day period itself. That such a drop hasn’t taken place yet indicates that stricter enforcement of social distancing guidelines is required throughout India. It is prudent to mention here that a significant majority of cases which the forecasts are based on, were detected during the lockdown period; an easing of the lockdown after 17 May is likely to lead to a spike in cases and a delay in the peak number of cases being reached. Especially for states such as Maharashtra and Delhi, where the peak is forecast to be reached after 17 May, it would be recommended that restrictions stay in place until a fall in cases is begun to be observed.

As with any study involving mathematical modeling, the current work involves caveats and limitations. The primary elements of the study are the data, the mathematical models applied and the numerical code, the sources and explanations for all of which have been included. However, future incidences such as the easing of social distancing measures may lead to a further increase in the number of cases than forecast. Additionally, factors that might increase the number of cases such as testing capacity have not been considered. While using the current forecast for a qualitative and quantitative understanding of the progress of COVID-19 in India, it would hence be prudent to consider these, as well as a number of other economics, social, demographic and medical factors.

## 4 Conclusion

The present work calibrated the Susceptible-Infected-Recovered (SIR) model and the Logistic Growth model to forecast the spread of SARS-CoV-2 virus using the data available till 13 May 2020 in India and its highest impacted states, namely Delhi, Maharashtra and Gujarat. The following are the primary conclusions from the same.

- The total number of cases forecasted using the SIR model were greater than those for the Logistic Growth model for all cases considered. The expected endpoints also show a similar trend, with the Logistic Growth model predicting an earlier endpoint in contrast with the SIR model for all regions.
- The endpoint of COVID-19 in India using the SIR model is predicted to be 12 September 2020. The Logistic Growth model forecasts the endpoint to be 23 July 2020.
- For the state of Maharashtra, the end point is expected to be 20 August 2020 as per the SIR model and 5 July 2020 as per the Logistic Growth model.
- Cases in Delhi are expected to converge on 9 September 2020 according to the SIR model and on 10 August 2020 according to the Logistic Growth model.
- The SIR model predicts an endpoint of 30 July 2020 for the state of Gujarat, while the Logistic Growth model forecasts the same to be 23 June 2020.
- Modelling the spread of COVID-19 using two distinct mathematical models has allowed us to tune different parameters derived from actual data, and has provided broad insights over the present situation.
- Our projections also take into account the various lockdowns and restrictive measures imposed by the Government of India. Analysing the data from various states has enabled us to identify changes in the trajectories of the spread of SARS-CoV-2 as a result of social distancing.
- It is recommended that current restrictive measures stay in place with even improved implementation in order that an eventual rise in cases doesn’t exceed the forecast values.

## Data Availability

No relevant data has been included.

